# Is migraine associated to brain anatomical alterations? New data and coordinate-based meta-analysis

**DOI:** 10.1101/2020.02.18.20024554

**Authors:** Rémy Masson, Geneviève Demarquay, David Meunier, Yohana Lévêque, Salem Hannoun, Aurélie Bidet-Caulet, Anne Caclin

## Abstract

A growing number of studies investigate brain anatomy in migraine using voxel-(VBM) and surface-based morphometry (SBM), as well as diffusion tensor imaging (DTI). The purpose of this article is to identify consistent patterns of anatomical alterations associated with migraine. First, 19 migraineurs without aura and 19 healthy participants were included in a brain imaging study. T1-weighted MRIs and DTI sequences were acquired and analyzed using VBM, SBM and tract-based spatial statistics. No significant alterations of gray matter (GM) volume, cortical thickness, cortical gyrification, sulcus depth and white-matter tract integrity could be observed. However, migraineurs displayed decreased white matter (WM) volume in the left superior longitudinal fasciculus. Second, a systematic review of the literature employing VBM, SBM and DTI was conducted to investigate brain anatomy in migraine. Meta-analysis was performed using Seed-based d Mapping via permutation of subject images (SDM-PSI) on GM volume, WM volume and cortical thickness data. Alterations of GM volume, WM volume, cortical thickness or white-matter tract integrity were reported in 72%, 50%, 56% and 33% of published studies respectively. Spatial distribution and direction of the disclosed effects were highly inconsistent across studies. The SDM-PSI analysis revealed neither significant decrease nor significant increase of GM volume, WM volume or cortical thickness in migraine. Overall there is to this day no strong evidence of specific brain anatomical alterations reliably associated to migraine. Possible explanations of this conflicting literature are discussed.

**Trial registration number:** NCT02791997, registrated February 6^th^, 2015.

## 1. Introduction

Migraine is the most common neurological disorder in the adult population with a prevalence comprised between 8% and 17% (Henry et al. 2002). Migraine attacks are characterized by acute, moderate to severe, recurrent headaches lasting between four to 72 hours, accompanied with nausea and/or hypersensitivity to visual (photophobia), auditory (phonophobia), olfactory (osmophobia) and/or tactile (allodynia) environmental stimulations (Headache Classification Committee of the International Headache Society (IHS) 2013). Several migraine subtypes (not necessarily exclusive) have been defined based on migraine attack frequency (episodic and chronic migraine), presence of aura preceding the attack (migraine with and without aura), or even secondary symptoms such as vertigo/head dizziness (vestibular migraine).

Neuroimaging methods have widely improved since the last two decades, especially through the popularization of automated whole-brain morphometric techniques such as voxel-(VBM) and surface-based morphometry (SBM) to assess gray- and white-matter volume from anatomical MRI images and tract-based spatial statistics (TBSS) to assess white matter microstructure from diffusion tensor imaging (DTI) data. These techniques enable to perform an unbiased investigation of structural changes in the brain associated with a condition, often without the need of precise a priori hypotheses. Morphometric analyses can reveal the “anatomical signature” of a disorder and therefore provide insights on the symptomatology of a disease. VBM is able to detect gray matter atrophy associated with Alzheimer’s disease (Frisoni et al. 2002; Karas et al. 2003). In various neuropsychological disorders such as excessive impulsivity (Matsuo et al. 2009), obsessive compulsive disorder (Valente et al. 2005), mild cognitive impairment (Chételat et al. 2002), attention-deficit and hyperactivity disorder (Makris et al. 2007), or dyslexia (Silani et al. 2005), gray matter abnormalities detected by VBM colocalizes with the brain network that was evidenced to be functionally altered in the disorder. As for DTI, it is a powerful tool to investigate the integrity of white matter fascicles and therefore to pinpoint potential abnormalities in brain connectivity in neurological disorders (Lim and Helpern 2002).

Identifying brain abnormalities associated with migraine is expected to provide new insights into the pathophysiology of the disease. The core symptoms of migraine may either alter brain structure through plastic mechanisms, or on the contrary originate from pre-existing brain abnormalities. It has been hypothesized that recurrent headaches over years may affect pain-related areas including somatosensory cortices or even lead to brain damage (May 2009). One can also expect that strategies used by migraine to cope with the symptoms (such as trigger avoidance or pain management) may have a lasting impact on brain structure. Indeed, structural alterations of the brain also reflect the effects of long-term plasticity as evidenced by the structural changes of gray matter volume in task-relevant brain regions following exercise and learning (Draganski et al. 2004; Boyke et al. 2008) or long-term practice of an instrument (Bermudez and Zatorre 2005; Bermudez et al. 2009). To respond to these questions, a rich and still growing literature have investigated brain anatomy in migraine using automatic morphometry techniques. Brain structural alterations in migraine have been reported by a great number of studies (May 2009; Bashir et al. 2013; Hu et al. 2015; Dai et al. 2015). However, results are often conflicting and no consensus has yet emerged identifying a structural signature of the disease. Previous meta-analyses on the subject have attempted to compile results from the literature and their conclusions are also inconsistent: some detected decreased gray matter volume in pain-related areas (Dai et al. 2015; Jia and Yu 2017), some in the frontal and cingulate cortices (Hu et al. 2015; Jia and Yu 2017) but the most recent one failed to detect any alteration of gray matter in migraine (Sheng et al. 2020).

The purpose of this article is to find out if there is convincing evidence of structural brain alterations associated to migraine. In order to respond to this question, we first provide new data aiming to identify gray (GM) and white matter (WM) abnormalities in patients with migraine during the interictal period. We aimed to provide a full picture of the migraine brain anatomy using several hypothesis-free whole-brain morphometry analyses in order to detect difference in gray and white matter volume (VBM), cortical thickness and shape (SBM), and the structural integrity of the white matter fascicles (TBSS). To anticipate, contrarily to our expectations based on previous reports, hardly any differences were found between the two groups in these analyses. Second, a systematical review of the literature was performed on whole-brain studies of GM and WM abnormalities in migraine in order to try to make sense of the conflicting results. To this end a coordinate-based meta-analysis (CBMA) of the literature was run using Seed-based d mapping via permutation of subject images (SDM-PSI), a novel meta-analysis technique which enables to detect robust and consistent structural alterations based on reported foci from different experiments (Radua et al. 2010; Albajes-Eizagirre et al. 2019b).

## 2. New data

### 2.1. Methods

#### 2.1.1. Participants

Twenty-five subjects were identified and diagnosed as migraineurs without aura by a neurologist specialized in cephalgia (GD, Hospices Civils de Lyon). Patients between 18 to 60 years old and reporting a migraine frequency between two to five attacks per month were included in this study. Exclusion criteria comprised migraine with aura, chronic migraine, a medical history of psychiatric or neurological disorders, ongoing background medical treatment other than contraceptive medication, and pregnancy. Patients who suffered from a migraine attack 72 hours prior to the scheduled MRI examination, were rescheduled at a later time, whereas those who suffered from a migraine attack within 72 hours post-MRI (n=6) were discarded from further analyses. Data from 19 migraineurs were thus retained for analyses (13 females, 6 males, mean age ± SD: 32.7 ± 8.7 years, all right-handed). Migraine patients filled the Headache Impact Test (HIT-6), a short questionnaire aiming to evaluate headache impact on everyday life (Kosinski et al. 2003) and the Migraine Disability Assessment Questionnaire (MIDAS) (Stewart et al. 1999) (Table 1).

**Table 1.** New Data. Demographics and headache profile of the control and migraine groups. HIT-6 scores are comprised between 36 (negligible impact of migraine on daily life) and 78. MIDAS scores between 0 and 5 correspond to little to no disability due to migraine, while scores higher than 21 correspond to a severe disability. Mean and standard deviation are provided when relevant. Group differences are tested using non-parametric Mann-Whitney U tests. NA: not applicable.

|  | Control | Migraine | p-value |
| --- | --- | --- | --- |
| **Sample size** | 19 | 19 | - |
| **Age (years)** | 33.6 (11.5) | 32.7 (8.7) | 0,93 |
| **Sex (number of female participants)** | 13 (68%) | 13 (68%) | - |
| **Total intracranial volume (mm3)** | 1543.1 (102.6) | 1546.9 (173.8) | 0,73 |
| **Laterality (number of right-handed)** | 19 | 19 | - |
| **Attacks per month** | NA | 3.3 (1.1) | - |
| **Migraine duration (years)** | NA | 16.8 (7.4) | - |
| **HIT-6 score** | NA | 64.2 (7.1) | - |
| **MIDAS score** | NA | 12.8 (12.1) | - |

Nineteen control subjects with no medical history of psychological or neurological disorders were identified from a cohort of sixty-three healthy participants for whom MRI scan were available and acquired with a procedure strictly identical to the migraine group. The MatchIt R package (Ho et al. 2011) was used to select subjects matched for age, sex and total intracranial volume (TIV), as those three covariates are known to have distinct contribution to GM volume (Pell et al. 2008). Propensity score matching was conducted using the nearest neighbor method (Caliendo and Kopeinig 2008) in order to minimize bias due to confounding factors. Additional demographic details are presented in Table 1. All persons gave their informed consent prior to their inclusion in the study.

#### 2.1.2. MRI acquisition

MRI examinations for all participants were performed on a Magnetom Prisma Siemens 3T MRI scanner equipped with a 64-channel head/neck coil. A T1-weighted sagittal magnetization-prepared-rapid acquisition with gradient echo (MPRAGE) image (repetition time (TR) = 3500 ms, echo time (TE) = 2.25 ms, inversion time (TI) = 1000 ms, field of view (FOV) = 250×250 mm, matrix size = 288×288, spatial resolution: 0.9×0.9×0.9 mm), and a diffusion tensor imaging (DTI) sequence with 64 gradient directions and 38 continuous slices (b = 0 and 1000 s/mm^2^, TR = 10000 ms, TE = 72 ms, FOV = 240×240 mm, matrix size = 132×132, spatial resolution: 1.8×1.8×1.8 mm) were acquired.

#### 2.1.3. Voxel-based and surface-based morphometry

The VBM and SBM analysis were conducted using the Computational Anatomy Toolbox (CAT12, dbm.neuro.uni-jena.de/cat/), an extension toolbox of Statistical Parametric Mapping software (SPM12, www.fil.ion.ucl.ac.uk/spm/software/spm12/). Default settings as detailed in the CAT12 manual (http://dbm.neuro.uni-jena.de/cat12/CAT12-Manual.pdf) were applied.

For the VBM analysis, GM, WM and cerebrospinal fluid (CSF) tissue segmentation was first performed. The resulting GM and WM masks were then aligned to the SPM12 tissue probability maps (DARTEL template in the MNI space), co-registered using DARTEL (Ashburner 2007) and smoothed with a Full-Width at Half-Maximum (FWHM) kernel of 10 mm (Ashburner 2015). TIV was finally estimated for each participant.

For the SBM analysis, the automated workflow from the CAT12 toolbox which measures cortical thickness and reconstructs the central surface was used. Based on the central surface data, we estimated the gyrification index and sulcus depth (Luders et al. 2006). Cortical thickness, gyrification index, and sulcus depth data were smoothed using a Gaussian filter with a FWHM kernel of 15 mm (default parameter).

#### 2.1.4. Statistical analyses (VBM and SBM)

Statistical designs were prepared using SPM12. A two-sample t-test was performed at each voxel to evaluate differences between migraineurs and control participants in regional GM and WM volumes, cortical thickness, gyrification index and sulcus depth data. For volumetric data, TIV, age and sex were considered as nuisance parameters and consequently entered in the design matrix as covariates as recommended in (Pell et al. 2008; Ashburner 2015). For surface data, only age and sex were entered as covariates. An implicit mask and a threshold masking (value=0.1) were applied to the images to remove voxels of the background from the analyses. Two contrasts were investigated: control > migraine, and migraine > control. Statistical inferences were made using non-parametric permutations and a Threshold-Free Cluster Enhancement (TFCE) correction was applied to the t-stats map produced (TFCE toolbox by Christian Gaser, http://dbm.neuro.uni-jena.de/tfce) to increase sensitivity (5000 permutations) (Smith and Nichols 2009) along with a family-wise error (FWE) correction to address multiple testing. Clusters were considered significant with p<0.05.

#### 2.1.5. DTI analysis

DTI analysis was conducted using tools from the FMRIB software library (FSL v5.0, fsl.fmrib.ox.ac.uk/fsl/fslwiki) and the Nipype pipelines (Gorgolewski et al. 2011). DTI images were first corrected for any susceptibility induced distortions (Smith et al. 2004), eddy currents, and head movements (Andersson and Sotiropoulos 2016). Fractional anisotropy (FA), mean diffusivity (MD), axial (AD) and radial diffusivity (RD) maps were calculated by fitting a tensor model at each voxel of the diffusion data. Tract-based spatial statistics (TBSS) was then performed on all participants’ FA images. TBSS analysis consisted first on non-linearly co-registering all FA images to the FMRIB58-FA standard-space template. A mean FA image was then created along with a skeleton of the major WM fiber tracts. Next, all participants FA images were projected onto this skeleton which will be fed into the voxelwise statistical analysis. This process was similarly applied to the MD, AD, and RD maps.

#### 2.1.6. Statistical analyses (DTI)

The voxel-level non-parametric permutation test (randomise function in FSL) was used to investigate the following contrasts on FA, MD, AD and RD maps: control > migraine and migraine > control. A TFCE correction was applied to the produced t-stats images to increase sensitivity (5000 permutations) along with a FWE correction to address multiple testing. Surviving clusters are reported with p<0.05.

#### 2.1.7. Power analysis

We ran a sensitivity power analysis using the G*Power software (Faul et al. 2007), using a power of 0.8 and an α error of 0.05 for our group comparisons. The required effect size given our sample size equals 0.82, suggesting an adequate sensitivity to large effects (Cohen 1977). Please note that it is challenging to perform power calculations for statistical analyses that involve high-dimensional data (Durnez et al. 2016), here power analysis was calculated as if we performed a simple one-tailed t-test. In neuroimaging studies, it is generally considered that a sample size above 16 participants is sensitive enough to detect medium and large effects most of the time (Friston 2012).

### 2.2. Results

#### 2.2.1. VBM results

Neither significant decreases nor significant increases of GM volume in migraine were detected. Compared to controls, WM volume was significantly decreased in migraine (corrected p=0.042, η^2^ = 0.78) in the left hemisphere which intersected the superior longitudinal fasciculus and the superior corona radiata (MNI coordinates of cluster peaks: [-30,-30,39], [-33,-26,26] and [-20,-38,39]), close to the superior temporal areas and the postcentral gyrus (Fig. 1). The reported effect size of this test is close to the required effect size of 0.82 as calculated by the G*Power software. No significant increases of WM volume in migraine were detected.

**Fig. 1.**
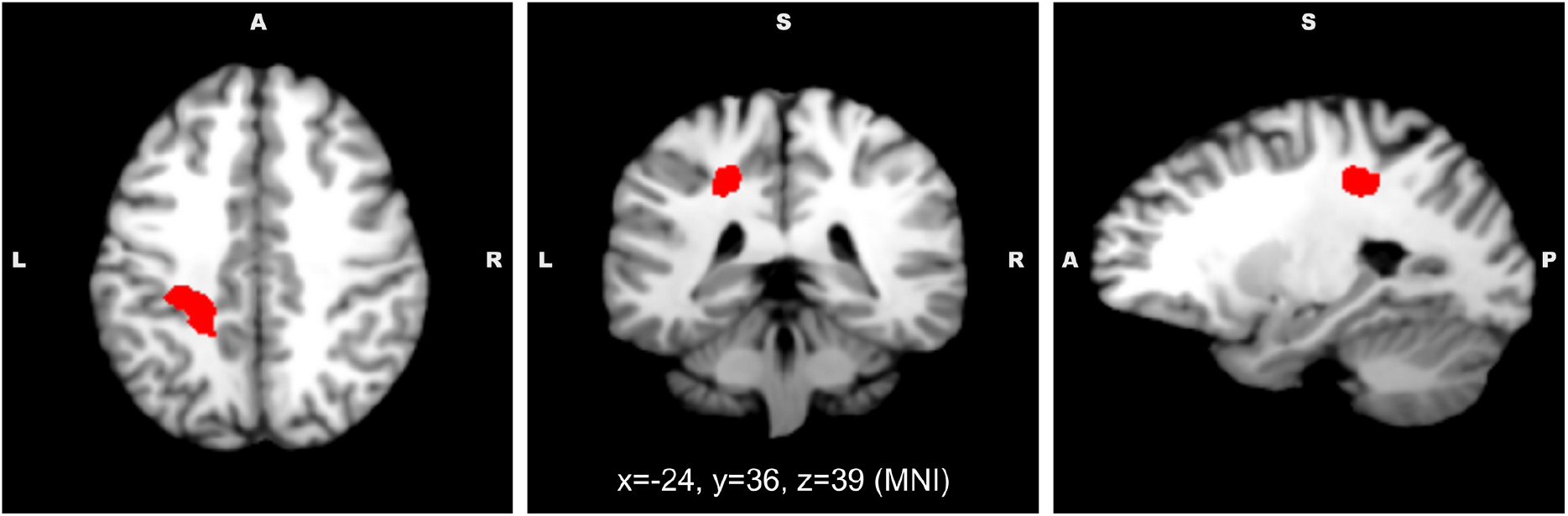
New Data. Voxels with a significant decrease of white matter volume in migraine participants (n=19) compared to healthy controls (n=19). From left to right, sagittal, coronal and axial views, MNI coordinates of the views are reported on the figure.

#### 2.2.2. SBM results

No significant differences in cortical thickness, gyrification index, and sulcus depth were detected between the control and migraine participants.

#### 2.2.3. DTI results

No significant differences in FA, MD, AD, or RD were detected between the control and migraine participants.

### 2.3. Discussion

With the present dataset, no brain anatomical differences could be detected in migraine regarding GM volume as assessed with VBM, cortical surface (thickness, gyrification and sulcus depth) as assessed with SBM, and in the integrity of WM as assessed with TBSS.

However, WM volume appeared to be decreased in migraine in the left superior longitudinal fasciculus (SLF). Only three studies (from the same research team) have yet reported WM volume decreases in migraine but none of them pinpointed such an alteration in the left superior longitudinal fasciculus (Schmitz et al. 2008; Arkink et al. 2017; Palm-Meinders et al. 2017) Moreover, no DTI study including the present one has reported altered WM integrity in this particular tract.

The SLF is an association tract which connects occipital and temporal areas to the frontal lobe. This pathway is involved in various cognitive processes (Schmahmann et al. 2008), however the left SLF has been consistently linked to language processing (e.g. Frye et al. 2010; Maldonado et al. 2011; Nagae et al. 2012; Madhavan et al. 2014) as it connects Broca’s and Wernicke’s area (Catani and Mesulam 2008). To our knowledge, migraine is not associated to major language defects, however some neuropsychological studies have reported that migraineurs performed worse than healthy participants in verbal memory and verbal skills tasks (for a systematic review, see Vuralli et al. 2018). The SLF might also be involved in the control of the vestibular function (Spena et al. 2006): an alteration of its integrity could underlie the vestibular symptoms observed in migraine as migraineurs are much more prone to vertigo and dizziness episodes than the general population (Vuković et al. 2007; Cha et al. 2009).

Overall we found little evidence for brain anatomical alterations in migraine. These negative results contrast with many studies in the literature which suggested that migraine was associated with abnormal brain structure and even a couple of meta-analyses which focused on gray matter volume (Dai et al. 2015; Hu et al. 2015; Jia and Yu 2017). In order to understand if our study is an outlier in the literature about migraine anatomy, we will now consider these negative results in light of a systematic review and a meta-analysis of previous studies on the subject.

## 3. Meta-analysis

### 3.1. Material and methods

#### 3.1.1. Data sources and study selection

Systematic searches were performed on January 2019 in PubMed database without any publishing time restriction. For VBM studies, we used the combination of keywords migraine AND ((voxel based morphometry) OR VBM); for SBM studies, we used the combination of keywords migraine AND ((surface based morphometry) OR (cortical thickness) OR (gyrification)); for DTI studies, we used the combination of keywords migraine AND ((diffusion tensor imaging) OR DTI). Additional studies were also searched from reference lists of the included articles. Inclusion criteria were: (1) the article was an experimental article; (2) it was published in an English-speaking peer-reviewed journal, (3) it included a VBM (gray and/or white matter), SBM, or DTI comparison of adult patients with migraine vs. healthy controls. If patient group overlapped with another study, the study with the larger sample size was retained. A paper was excluded if the patient group was not afflicted with migraine as defined by the International Headache Society (Headache Classification Committee of the International Headache Society (IHS) 2013) (presence of cluster headache, medication-overuse headache, tension headache, etc.). Furthermore, studies performing whole-brain analysis and reporting results coordinates in a standard stereotactic space (MNI or Talairach) were separated from studies performing ROI analysis or that failed to include stereotactic coordinates of the results. For DTI studies, we did not exclude articles which did not report stereotactic coordinates of the results as it did not appear to be a common practice. For each paper, demographics and headache profile of the sample and analysis methods were extracted.

#### 3.1.2. SDM-PSI meta-analysis

Regarding VBM or SBM studies, if a sufficient number of studies (>10 studies) was obtained during the systematic review of the literature, results of those studies were combined with Seed-based d Mapping with Permutation of Subject Images (SDM-PSI) using the SDM-PSI software (version 6,21, sdmproject.com) to identify brain structures that were consistently affected in migraine. The full SDM-PSI procedure is described in the tutorial available on their website (Albajes-Eizagirre et al. 2019a). First, when a study reported a significant difference between the control and migraine group, coordinates of the peaks of significant clusters and the associated t-value were extracted. If the original study reported *z-s*cores or p-values instead of *t-values*, they were converted to t-values using the online tool provided by the SDM project website (sdmproject.com). Negative studies were included in the meta-analysis. Preprocessing was performed according to standard SDM-PSI parameters, using a 20 mm full width half maximum (FWHM) anisotropic Gaussian kernel and 2mm voxel size. As advocated by SDM-PSI guidelines, significant results were thresholded using a family-wise error (FWE) correction based on threshold-free cluster threshold enhancement (TFCE) with corrected p-value threshold of 0.05 and a minimal cluster extent of 10 voxels. However, this FWE correction can be an overly conservative strategy in some situations and lead to false negative results (Albajes-Eizagirre et al. 2019b). Consequently, significant results were also thresholded with an uncorrected p-value threshold of 0.005 and a minimal cluster extent of 10 voxels, a statistical strategy which has been considered to provide an optimal balance between sensitivity and sensibility (Radua et al. 2012).

### 3.2. Results

#### 3.2.1. VBM – Gray matter

The search strategy resulted in 61 relevant documents among which only 23 were retained (Table 2, Figure 2). Some articles investigated more than one subtype of migraine: results are then considered separately for the meta-analysis elevating the number of “actual” studies to 32. They involved a total of 1172 healthy participants and 1071 migraineurs.

**Table 2.**
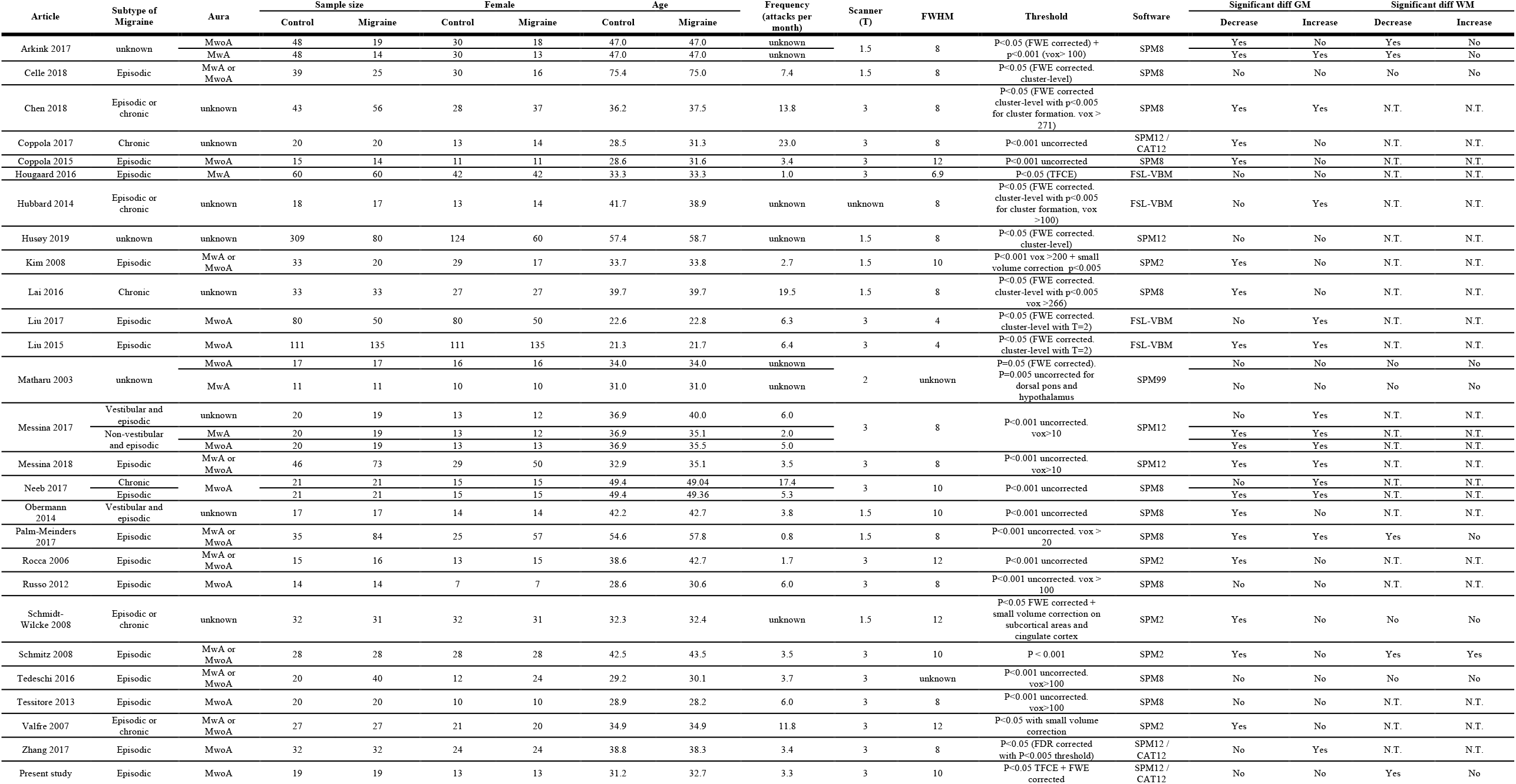
Summary of the voxel-based morphometry (VBM) studies included in the meta-analysis. N.T. = not tested, MwA = migraine with aura, MwoA = migraine without aura, FWE = family-wise error, FDR = false detection rate, TFCE = threshold-free cluster enhancement, vox = voxels, FWHM = full-width height maximum.

**Fig. 2.**
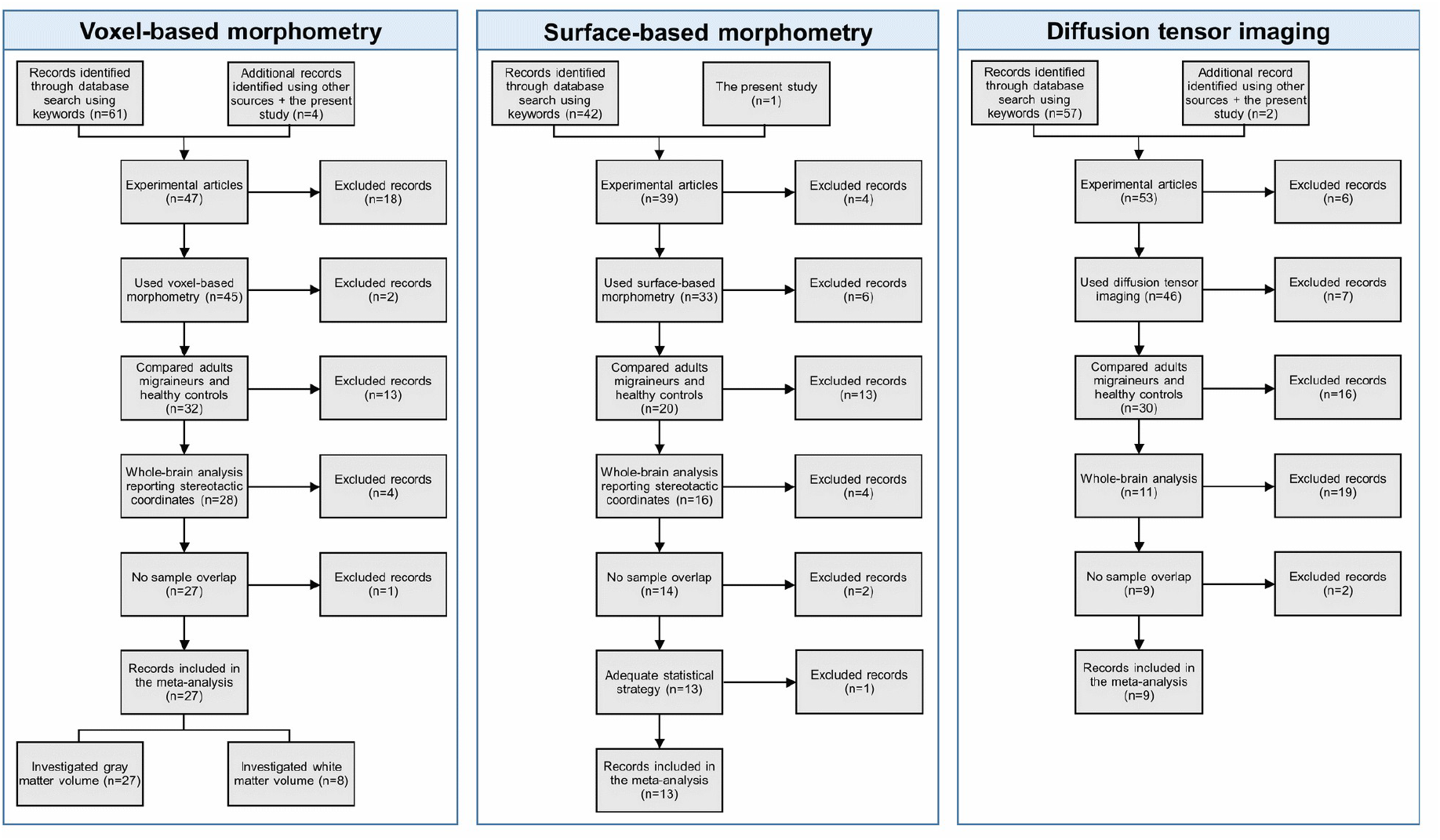
Search strategy used for the inclusion of the studies considered in the present meta-analysis.

Out of 32 studies, 23 (72%) found differences in GM volume in migraine. 18 (56%) observed a decrease and 13 (41%) an increase of GM volume in the brain, including 8 studies (25%) reporting both GM volume increases and decreases in migraine. The meta-analysis indicated no consistent GM volume increase or decrease in the migraine group, even with the more lenient statistical strategy (uncorrected p<0.005).

#### 3.2.2. VBM – White matter

Out of the 27 articles retained for the meta-analysis of GM volume, only 8 analyzed WM volume. 2 of these eight investigated two subtypes of migraine elevating the number of “actual” studies to 10 (Table 2, Figure 3). They involved a total of 249 healthy participants and 269 migraineurs.

**Fig. 3.**
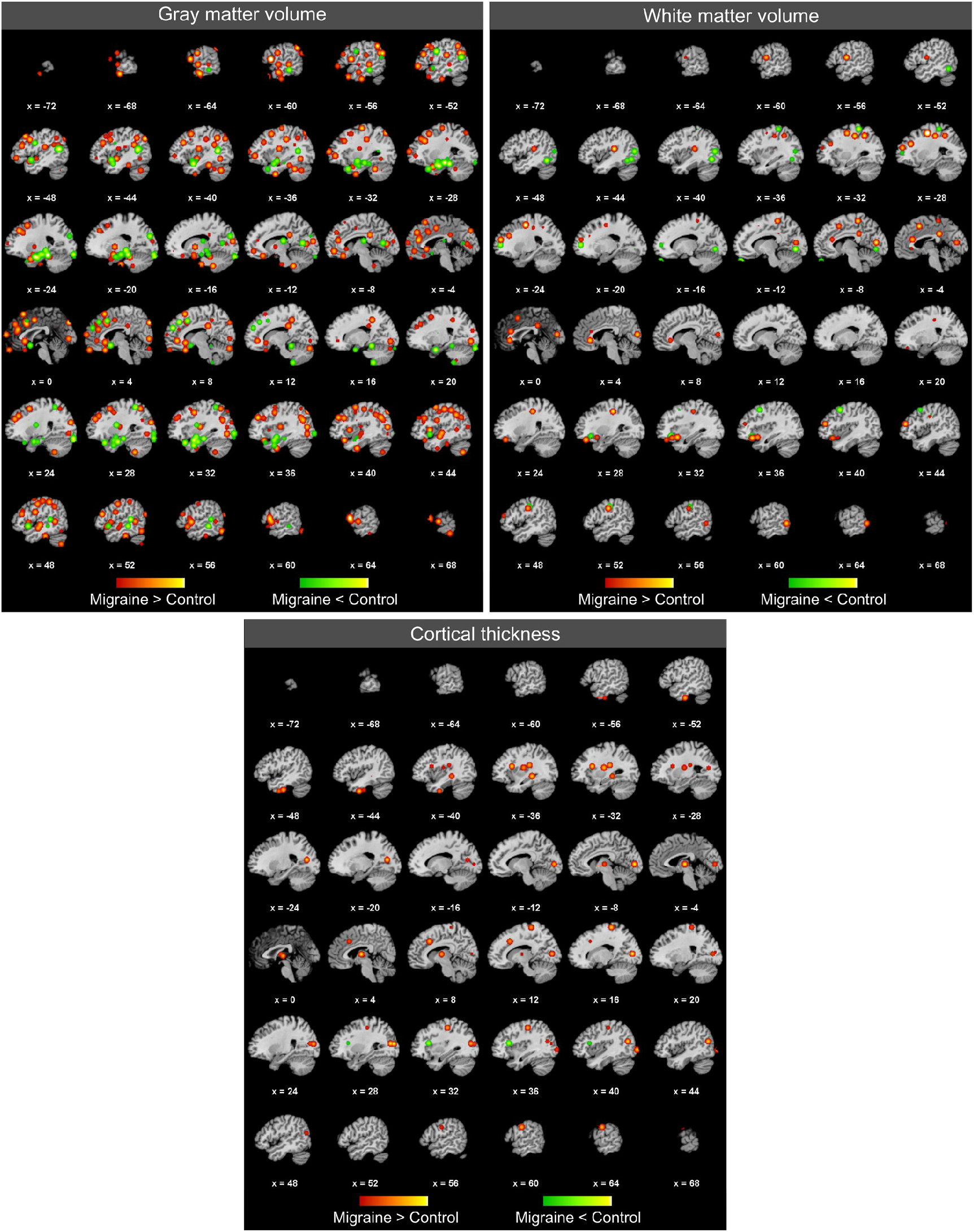
Here are reported on a standard T1-weighted image all loci from the literature in which a significant difference in grey matter volume, white matter volume and cortical thickness in migraine has been detected (respectively n=32, n=10 and n=16 studies). Foci are blurred using a Gaussian filter with a Full-Width Height Maximum value computed based on the sample size of the study. The green gradient corresponds to an increase in migraine, the red gradient corresponds to a decrease in migraine. Please note that some studies have reported only the peak of significant clusters while others also reported also local maxima inside the significant cluster: a relatively high concentration of foci may not necessarily reflect convergence between studies.

Out of ten studies, five (50%) found increases in WM volume in migraine, including one study (10%) reporting both WM volume increases and decreases in migraine. The meta-analysis indicated no consistent WM volume increase or decrease in the migraine group, even with the more lenient statistical strategy (uncorrected p<0.005).

#### 3.2.3. SBM – Cortical thickness

The search strategy resulted in 42 relevant documents among which only 12 were retained (Table 3, Fig. 2). Some articles investigated two subtypes of migraine: results are then considered separately for the meta-analysis elevating the number of “actual” studies to 16 (Table 3). They involved a total of 848 healthy participants and 776 migraineurs.

**Table 3.** Summary of the surface-based morphometry (SBM) studies included in the meta-analysis. N.T. = not tested, MwA = migraine with aura, MwoA = migraine without aura, FWE = family-wise error, FDR = false detection rate, TFCE = threshold-free cluster enhancement, vox = voxels, FWHM = full-width height maximum.

|  | Subtype of Migraine | Aura | Sample size |  | Female |  | Age |  | Frequency (attacks per month) | Scanner (T) | Threshold | Software | Cortical thickness |  | Cortical gyrification |  | Sulcus depth |  |
| --- | --- | --- | --- | --- | --- | --- | --- | --- | --- | --- | --- | --- | --- | --- | --- | --- | --- | --- |
|  |  |  | Control | Migraine | Control | Migraine | Control | Migraine |  |  |  |  | Decrease | Increase | Decrease | Increase | Decrease | Increase |
| Datta 2011 | Episodic | MwoA | 28 | 28 | 24 | 24 | 33.0 | 35.0 | 3.4 | 3 | P<0.05 (permutations + FDR corrected) | Freesurfer | No | No | N.T. | N.T. | N.T. | N.T. |
|  |  | MwA | 28 | 28 | 24 | 24 | 33.0 | 35.0 | 4.1 |  |  |  | No | No | N.T. | N.T. | N.T. | N.T. |
| Gaist 2018 | Episodic or chronic | MwA | 137 | 166 | 137 | 166 | 48.0 | 48.0 | unknown | 3 | P<0.05 (Monte-Carlo) | Freesurfer | No | Yes | N.T. | N.T. | N.T. | N.T. |
| Hougaard 2016 | Episodic | MwA | 60 | 60 | 42 | 42 | 33.3 | 33.3 | 1.0 | 3 | P<0.05 (Cluster-based permutations) | FSL-VBM | No | No | N.T. | N.T. | N.T. | N.T. |
| Hubbard 2014 | Episodic or chronic | unknown | 18 | 17 | 13 | 14 | 41.7 | 38.9 | unknown | unknown | P<0.005 (Cluster forming threshold p<0.005, vox > 100, random-field-theory-based corrected) | FSL-VBM | Yes | No | N.T. | N.T. | N.T. | N.T. |
| Husøy 2019 | Episodic or chronic | unknown | 309 | 80 | 124 | 60 | 57.4 | 58.7 | unknown | 1.5 | P<0.05 (FDR) | Freesurfer | No | No | N.T. | N.T. | N.T. | N.T. |
| Kim 2014 | Episodic | MwoA | 34 | 56 | 56 | 34 | 34.2 | 35.7 | 2.5 | 3 | P<0.05 (Monte-Carlo) | Freesurfer | No | Yes | N.T. | N.T. | N.T. | N.T. |
| Maleki 2015 | Episodic or chronic | unknown | 46 | 46 | 46 | 46 | 34.1 | 34.7 | unknown | 3 | P<0.05 (Monte-Carlo) | Freesurfer | No | Yes | N.T. | N.T. | N.T. | N.T. |
| Messina 2013 | Episodic or chronic | MwoA | 18 | 31 | 13 | 22 | 37.2 | 38.6 | unknown | 3 | P<0.01 uncorrected vox > 100 / p<0.05 FDR | Freesurfer | Yes | No | N.T. | N.T. | N.T. | N.T. |
|  | Episodic or chronic | MwA |  | 32 | 13 | 20 | 37.2 | 35.2 | unknown |  |  |  | Yes | No | N.T. | N.T. | N.T. | N.T. |
| Petrusic 2018 | Episodic | MwA | 30 | 48 | 23 | 36 | 39.6 | 39.3 | 0.7 | 1.5 | P<0.05 (Monte-Carlo) | FSL 5 – Freesurfer | No | No | N.T. | N.T. | No | No |
| Zhang 2017 | Episodic | MwoA | 32 | 32 | 24 | 24 | 38.8 | 38.3 | 3.4 | 3 | P<0.05 (Cluster forming threshold p<0.005 + FDR corrected) | SPM12 / CAT12 | Yes | Yes | Yes | Yes | N.T. | N.T. |
| Woldeamanuel 2019 | Chronic | MwA or MwoA | 30 | 30 | 24 | 24 | 40.0 | 40.0 | 27.0 | 3 | P<0.001 + FDR corrected or cluster corrected | Freesurfer | No | No | N.T. | N.T. | N.T. | N.T. |
| Magon 2018 | Episodic | MwA | 115 | 38 | 81 | 109 | 29.1 | 30.8 | 3.3 | 3 | P<0.05 (FDR corrected) | Freesurfer | Yes | No | N.T. | N.T. | N.T. | N.T. |
|  |  | MwoA |  | 93 |  |  |  |  |  |  |  |  | Yes | No |  |  |  |  |
| Present study | Episodic | MwoA | 19 | 19 | 13 | 13 | 31.2 | 32.7 | 3.3 | 3 | P<0.05 TFCE + FWE corrected | SPM12 / CAT12 | No | No | No | No | No | No |

Out of 16 studies, 9 studies (56%) found differences in cortical thickness in migraine. 6 studies (38%) observed decreases of cortical thickness, 4 studies (25%) observed increases of cortical thickness and 1 study (6%) reported both increases and decreases of cortical thickness in migraine. The meta-analysis indicated no consistent cortical thickness increase or decrease in the migraine group, even with the more lenient statistical strategy (uncorrected p<0.005).

##### Others surface metrics

Only two studies (including the present one) have investigated cortical gyrification. Zhang and colleagues found an increased gyrification index in left postcentral gyrus, superior parietal lobule and right lateral occipital cortex, and decreased gyrification index in the left rostral middle frontal gyrus in migraine (Zhang et al. 2017), while our study did not observe any group difference. Only two studies (including the present one) have investigated sulcus depth and none of them detected a significant difference between migraine and control groups (Zhang et al. 2017).

#### 3.2.4. DTI

The search strategy resulted in 57 relevant documents among which only 7 were retained. Some articles investigated two subtypes of migraine: results are then considered separately for the meta-analysis elevating the number of “actual” studies to twelve (Table 4). They involved a total of 252 healthy participants and 352 migraineurs.

**Table 4.**
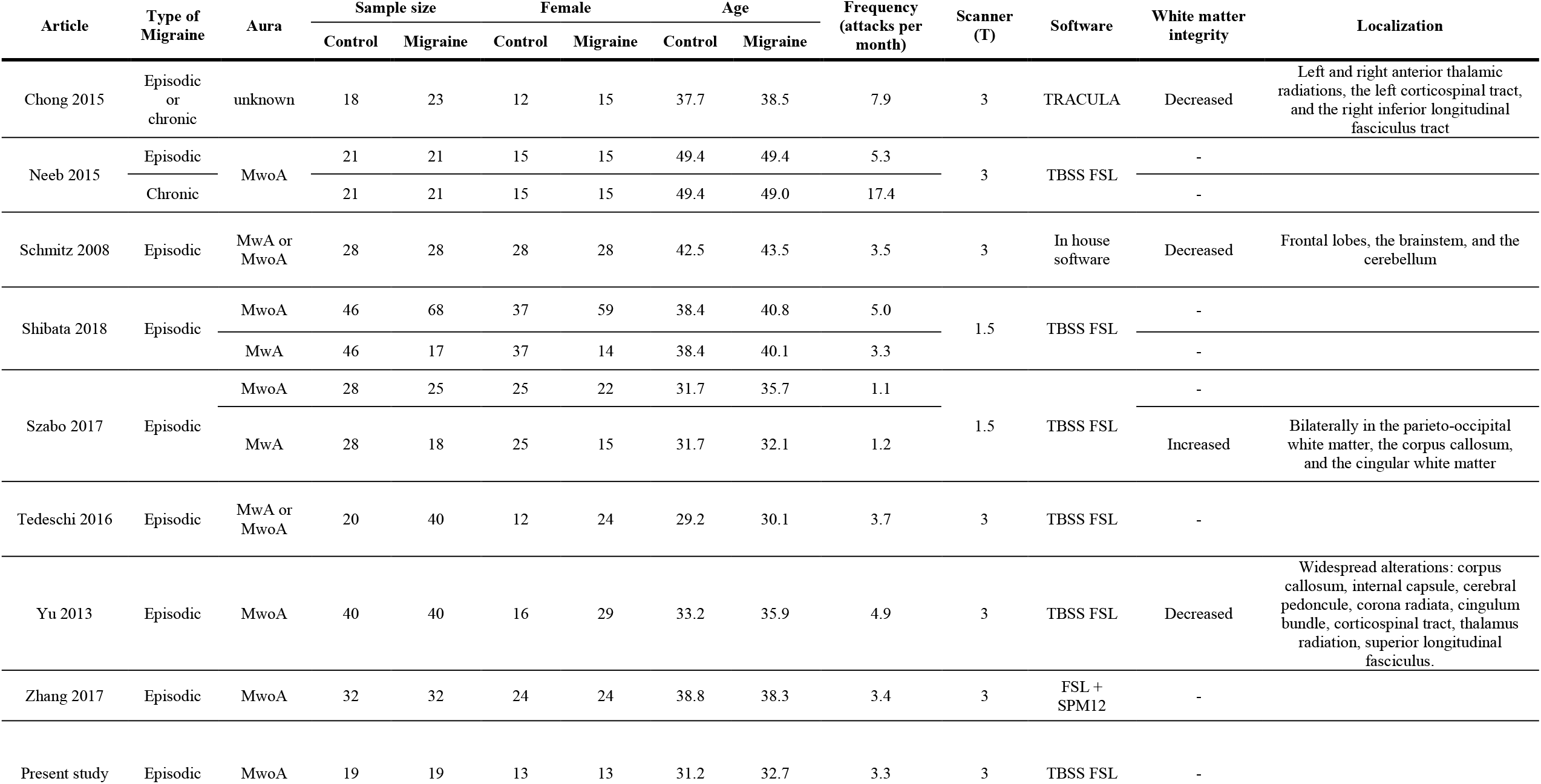
Summary of the diffusion tensor imaging studies (DTI) included in the meta-analysis. N.T. = not tested, MwA = migraine with aura, MwoA = migraine without aura, TBSS = tract-based spatial statistics.

No SDM-PSI analysis was conducted regarding DTI studies as no stereotactic coordinates of significant results were reported in the articles. In the following, we will consider *decreased* FA or AD and *increased* MD or RD as a sign of altered WM integrity. Out of twelve studies, four (33%) found differences in WM integrity. Decreased WM integrity in migraine was detected in three studies (25%), which involved 86 healthy participants and 91 migraineurs, in different fiber tracts depending on the studies. Only one study (8%) reported increased WM integrity in migraine. Further information is available in Table 4.

### 3.3. Discussion

All studies considered here investigated brain structures during the interictal period, for obvious practical considerations and also because some results suggest that the migraine headaches cause transient changes of the brain structure (Coppola et al. 2015), which may not reflect long-term alterations of the migraine brain. It is noteworthy that migraine symptoms are not exclusive to the ictal period. If sensory disturbances clearly climax during the attacks, alterations of sensory processing extend beyond the ictal state (Main et al. 1997; Vingen et al. 1998; Granovsky et al. 2018; Lévêque et al. 2020). Migraine may be associated to minor cognitive dysfunctions interictally (Zeitlin and Oddy 1984; Hooker and Raskin 1986; Mongini et al. 2005; Vuralli et al. 2018) and to vertigo and dizziness episodes (Vuković et al. 2007; Cha et al. 2009).

SDM-PSI analysis is a powerful tool to investigate the spatial convergence of reported structural alterations in morphometry studies. It is an improvement compared to previous coordinate-based meta-analysis (CBMA) methods such as the popular Activation Likelihood Estimation (ALE) (Eickhoff et al. 2012), notably because it allows to take into account studies with null results and the peaks’ effect size in positive studies, and uses the jack-knife analysis to limit the contribution of a small subset of studies. The systematic review of the literature resulted in a sufficient number of studies for a SDM-PSI analysis for GM volume, WM volume, and cortical thickness. For either of these metrics, no significant difference between the control and the migraine groups was detected, even with a lenient statistical threshold. Regarding WM volume, the relatively low number of studies makes any conclusion uncertain. It appears that there is a tendency of WM loss on migraine as half of the studies reported WM volume decrease while only one reported WM volume increase. However, reported loci of WM volume decrease are relatively scattered across the brain. The situation is even more obscure concerning cortical thickness, since a similar number of studies reported cortical thickness increase and decrease which affected cortical areas dispersed across the cortical surface.

Finally, regarding DTI, a minority of studies reported alteration of white matter tracts in migraine. When they did, reported anatomical alterations were generally widespread but did not necessarily intersected across studies. In studies only investigating regions of interest (not presenting whole-brain analyses), alterations of white matter integrity in migraine were reported in regions as diverse as the thalamus (Coppola et al. 2014), the brainstem (Kara et al. 2013; Marciszewski et al. 2018), the corpus callosum (Li et al. 2011; Yuan et al. 2012), visual processing networks (Granziera et al. 2006; Rocca et al. 2008) or fronto-insular tracts (Gomez-Beldarrain et al. 2016; Liu et al. 2018).

In conclusion, for these three metrics of brain anatomical integrity, there is no emerging pattern of anatomical alteration in migraine.

## 4. General discussion

The question which underlies this whole study was quite simple: *are there chronic anatomical alterations of the brain associated to migraine?* In spite of a rich and growing literature, we are still far from a consensus on whether migraineurs present such alterations and which brain areas are potentially affected. Previous studies reported highly heterogeneous results, either in terms of the presence of a group effect or in terms of the direction and the localization of a potential effect. Can we make sense of this conflicting literature?

### 4.1. Heterogeneity of protocols, heterogeneity of results?

As illustrated in the tables 2 to 4, there exists quite a heterogeneity in the protocols chosen in previous studies in the literature.

First, numerous studies have favored investigating one subtype of migraine (migraine with/without aura, episodic/chronic migraine, vestibular migraine), in an attempt to reduce the variability in the migraine group. Previous results suggest that migraineurs with aura may differ anatomically from migraineurs without aura in terms of GM volume (Messina et al. 2017), of cortical thickness (Magon et al. 2018), and white matter integrity (Szabó et al. 2017; Shibata et al. 2018), highlighting the importance of considering the two groups separately. Vestibular migraine differed from other types of migraine in terms of GM volume (Messina et al. 2017). Finally, GM damage appears to be increased in chronic compared to episodic migraine (Neeb et al. 2017; Chen et al. 2018) and it correlates with attack frequency (Valfrè et al. 2007; Kim et al. 2008; Schmitz et al. 2008; Neeb et al. 2017; Messina et al. 2018). In conclusion, based on the literature, it is probable that each subtype of migraine presents a specific anatomical signature. More studies are needed for this hypothesis to be tested in a meta-analysis.

Second, all the studies considered here are not necessarily homogenous in terms of demographic characteristics. The mean age of the migraine sample ranges from under 30 to over 70 years old while there are suspicions that anatomical alterations evolve with age (Schmitz et al. 2008; Liu et al. 2013; Chong et al. 2014; Neeb et al. 2017; Messina et al. 2018). Gender seems to interact with the pathology (Dai et al. 2015), yet some studies chose to only include women and other (including the present study) opted for a sex-ratio closer to the migraine sex-ratio in the general population. Other variables such as comorbidities, education level, or medication overuse may interact with the pathology and affect the patterns of anatomical alterations.

Finally, if voxel-based and surface-based morphometry analyses are based on standardized, streamlined workflows, slight deviations in the parameters can affect results in a major way. As illustrated in the tables 2 to 4, there are discrepancies on the statistical thresholds applied in such analyses: some studies have opted for uncorrected p-values, which is often an overly lax statistical strategy, or for a cluster-level control of FWE (implemented by default in SPM12 statistics) which is unlikely to be appropriate for VBM as it assumes stationary smoothness (Ridgway et al. 2008). Inappropriate or lax statistical strategies may have led to a disproportionate rate of false positives, accounting for some of the heterogeneity in previous results. However, if we presume that there is a major anatomical alteration in migraine (i.e. with a large effect size), it should have been detected consistently, irrespective of the statistical strategy and therefore it should have been revealed through this meta-analysis.

### 4.2. A lack of statistical power?

Small sample sizes can be appropriate for exploratory studies as trivial effects are very unlikely to reach significance which ensures that only large-sized effects with actual scientific importance will be detected (Friston 2012). However, low statistical power reduces the reproducibility of the results and increase the probability of false positives (Button et al. 2013). Moreover, if subtle effects are to be expected, scrupulous matching of the control participants is crucial in order to avoid the detection of spurious effects (May 2009).

Statistical power is usually relatively satisfactory in the studies considered in this meta-analysis. Most studies presented a sample size superior to 20 participants (in each group), especially in SBM and DTI studies. Some of them presented a sample size superior to 60 participants, ensuring the detection of even small effects and a limited probability of a false positive (Friston 2012). Interestingly, out of the three VBM studies with a large sample size (>60), only one of them detected an effect on GM volume. Out of the three SBM studies with a large sample size (>60), two of them detected an effect on cortical thickness, but not in the same direction. Such observations do not support the hypothesis of the presence of brain anatomical alterations in migraine.

### 4.3. The issue of publication bias

Publication bias is a widespread concern which is known to distort the results of meta-analyses as positive results are more likely to be published than negative results (Thornton and Lee 2000). This risk is consubstantial to any attempt of performing a meta-analysis. There exist tools to evaluate publication bias in coordinate-based meta-analyses based of the reported effect sizes of the significant loci (Acar et al. 2018). However, their role is to assess the robustness of convergent results, a prerequisite which is not fullfilled in the present situation. Beyond the fact that the reported structural alterations associated to migraine in the literature are widely inconsistent, there are other serious signs that the publication bias might be particularly exacerbated in the present situation.

First, anatomical images (especially T1-weighted MRI images) are routinely acquired in numerous studies, notably in functional studies using fMRI. It is very likely that many scientific teams have usable datasets available for morphometry analyses. Second, voxel- and surface-based morphometry are fairly simple to use and widely available techniques, as streamlined workflows exist in two common free analysis toolboxes (SPM and FSL). They do not necessitate much of computing power nor are they too time-consuming. It is reasonable to assume that numerous researchers in the field of migraine have attempted to analyze their anatomical data but that a large part of these analyses have never got published due to unconvincing results. Regarding VBM and SBM studies, even in the available literature, between one third and half of the articles did not report any significant difference between the control and migraine participants. It is probable that this proportion of negative results would be much higher if unpublished analyses were to be considered. Such proportions do not reassure on the actual presence of anatomical alterations in migraine.

This reasoning is not as appropriate for DTI studies as diffusion sequences are not routinely acquired in functional studies and as DTI analysis workflows are less common and streamlined than their VBM counterparts.

## 5. Conclusions and future directions

Previous studies reporting anatomical alterations in migraine do not converge neither on the direction nor on the spatial localization of the effect. Negative results are quite prevalent, especially in the context of a potentially strong publication bias. Based on current knowledge, there is to this day no strong evidence for the presence of systematic brain anatomical abnormalities associated to migraine. However, this study alone is not sufficient to rule out the existence of subtle anatomical alterations in migraine nor the existence of alterations specific to some migraine subtypes. Also, the number of studies on WM integrity and cortical surface in migraine is still quite low leading to weak conclusions. Further research is needed to produce a better picture.

What could be the next steps in researching brain anatomy alterations in migraine?

Small-sized, exploratory studies do not appear to be sufficient to shed light on possible anatomical alterations in migraine, especially regarding GM alterations. If a large-size effect existed, it should have been consistently reported by these studies. However, it remains scientifically crucial to keep on reporting morphometry analyses results, even if the statistical power is low, in order to provide information for future meta-analyses.

One of the major future developments could be longitudinal studies at different timescales. Migraine has been postulated to be a progressive disease with brain damage accumulating over the years, even if this proposition is controversial (May 2009). To our knowledge, at least two studies have attempted to study long-term effects of migraine (after a one-year or a four-year follow-up evaluation) with promising results (Liu et al. 2013; Messina et al. 2018). Further research is needed to confirm those results. It would be particularly interesting to investigate through longitudinal studies if spontaneous migraine remission with age is associated to a receding of anatomical alterations. On a different timescale, it has been suggested that anatomical alterations evolve along the migraine cycle (Coppola et al. 2015). All but one study in this article reported structural images during the interictal period. Deeper understanding of the dynamics of brain plasticity during the migraine cycle through short-term longitudinal studies would be of great interest.

## Data Availability

Data that support the findings are not publically available because participants included in this study did not consent to sharing their personal data with other parties.

## 6. Acknowledgments

The acquisition of imaging data was performed at the CERMEP imaging center in Lyon, we thank Frank Lamberton for his technical assistance. We thank Hesham ElShafei and Lesly Fornoni for their help in recruiting the participants.

## Declarations

### Ethics approval

The ethical approval of this work was obtained through the Hospices Civils de Lyon, approved by the local ethical committee (Comité de Protection des Personnes SUD EST III). Therefore, this work has been performed in accordance with the ethical standards laid down in the 1964 Declaration of Helsinki and its later amendments.

### Consent to participate

Written informed consent has been obtained from all participants in the present study.

### Consent to publish

Participants have signed written consent regarding publishing results derived from analyses of their data.

### Availability of data and material

The data that support the findings of this study are not publically available as participants did not consent to data sharing.

